# Comparative functional survival and equivalent annual cost of three long lasting insecticidal net (LLIN) products in Tanzania

**DOI:** 10.1101/19002212

**Authors:** Lena M Lorenz, John Bradley, Joshua Yukich, Dennis Joram Massue, Zawadi Mageni Mboma, Olivier Pigeon, Jason Moore, Albert Kilian, Jo Lines, William Kisinza, Hans J Overgaard, Sarah J Moore

## Abstract

Almost 1.2 billion long-lasting insecticidal nets (LLINs) have been procured for malaria control. Institutional buyers often assume that World Health Organization (WHO) prequalified LLINs are functionally identical with a three-year lifespan. We measured the lifespans of three LLIN products, and calculated their cost-per-year of functional life, through a randomised double-blinded prospective evaluation among 3,420 study households in Tanzania using WHO-recommended methods. Primary outcome was LLIN functional survival (LLINs present in serviceable condition). Secondary outcomes were 1) bioefficacy and chemical content (residual insecticidal activity) and 2) protective efficacy for volunteers sleeping under LLINs (bite reduction and mosquitoes killed). LLIN median functional survival was significantly different: 2·0 years for Olyset, 2·5 years for PermaNet and 2·6 years for NetProtect. Functional survival was affected by accumulation of holes resulting in users discarding nets. Protective efficacy also significantly differed between products as they aged. The longer-lived nets were 20% cheaper than the shorter-lived product.

## Introduction

Sleeping under LLINs remains the most cost-effective way to control malaria and reduce mortality,^1^ notwithstanding insecticide resistance.^2^ However, despite 254 million LLINs being procured globally in 2017 alone,^3^ global LLIN coverage remains inadequate, with only 56% of the population in endemic areas estimated to have access to a LLIN.^3^ LLINs are mostly distributed through periodic mass distribution campaigns, and as a result, population access to LLIN fluctuates over time. Access is typically high directly after a mass campaign and then drops as low as 50% just before the next campaign, as nets wear out. This insufficiency is particularly salient because gains in malaria control have stalled, with fewer than 50% of endemic countries remaining on track to reach critical malaria reduction targets.^3^ Investment in malaria control has stagnated and was US$1·3 billion (30%) below the resources required in 2017 to meet World Health Organization (WHO) targets of reducing malaria case incidence and mortality rates by at least 40% by 2020.^3^ These gaps in funding and coverage emphasise the need to deploy products that present the best value for money.

A report to the Malaria Policy Advisory Group (MPAC) advised that increasing the functional life of long lasting insecticidal nets (LLINs) by one or two years, would reduce the cost of malaria control by between US$500-700 million over five years.^4^ Currently, the WHO prequalifies LLIN products that demonstrate adequate insecticidal activity three years after deployment, but do not appraise the physical deterioration of nets over time as part of LLIN prequalification assessment. This has resulted in a tendering process where LLINs are assumed by donors to be identical, and procurement is weighted by the unit price of the commodity and not product lifespan.^5^ However, all the available data suggests that the assumption of uniform three-year lifespan for all LLIN products is unrealistic.^6^ There is a clear need for a more integrative economic approach to base purchasing decisions on value-for-money and cost-per-effective unit of LLIN coverage.^4,5^ New product classes of LLINs with novel active ingredients for insecticide resistance management are becoming available,^7^ but they remain susceptible to the same forces of physical disintegration, being discarded and losing insecticidal activity. Moreover, in most cases they cost more. This emphasises the need to consider price of LLINs in terms of cost per year of functional life.^8^

A functional LLIN is one that is present, in good physical condition and remains insecticidal, thereby providing protection against vector-borne diseases through preventing bites and killing disease vectors.^4^ Durability, or functional survival, of LLINs varies between geographical regions^9^ and environments^10,11^ and remains an under-valued, yet essential determinant of the success and efficiency of malaria control programmes.^12,13^ How long LLINs remain protective under user conditions will dictate how frequently they must be replaced, which has both public health and economic implications.^8^ In 2011, it was calculated that in Tanzania, for mean LLIN lifespan of two, three and four years, 89, 63 and 51 million LLINs, respectively, would be needed over ten years to achieve national access targets.^12^

Here we report results from a large prospective durability study of three LLIN products, conducted in Tanzania. The proportion of LLINs remaining in use and still protective against malaria mosquitoes was measured over three years follow-up after deployment. We calculated relative LLIN cost-effectiveness in terms of the equivalent annual cost (EAC), which is a conventional financial indicator used to compare products with different effective life-times. The median functional survival of each product and its EAC was calculated to inform optimal procurement of cost-effective LLINs.^12^

## Results

A total of 3,393 households were randomised to which 10,571 nets were distributed (3,520 Olyset (33%), 3,513 PermaNet 2.0 (33%) and 3,538 NetProtect (33%)). The three study arms were similar in number of participants, number of nets allocated, household characteristics, house design and socioeconomic characteristics (Table S1). Households lost to follow up was 20% over the three years of the trial.

### Functional Survival

There were significant differences between functional survival of the three products (defined as presence of serviceable nets) (Table 1). Estimated median functional survival was 2·0 years for Olyset, 2·5 years for PermaNet and 2·6 years for NetProtect. There was no significant difference in net use by net product (Table S2).

**Table 1:** Percentage net functional survival (defined as presence of the net in the house and in serviceable condition) with 95% confidence intervals in parenthesis and simulated Equivalent Annual Cost (assuming USD $3·0 purchase price) by net product and time point

| Net product | % Functional survival (95% CI) |  |  | Median survival in years (95% CI) | Hazard ratio (95% CI) | Simulated Equivalent Annual Cost in USD (95% CI) |
| --- | --- | --- | --- | --- | --- | --- |
|  | 10 months | 22 months | 36 months |  |  |  |
| Olyset | 82 (79, 85) | 54 (47, 62) | 27 (20, 34) | 2.0<br>(1.7, 2.3) | 1 | 1.5<br>(1.3, 1.7) |
| PermaNet | 88 (85, 90) | 65 (57, 72) | 38 (31, 46) | 2.5<br>(2.2, 2.8) | 0.73<br>(0.64, 0.85) | 1.2<br>(1.1, 1.4) |
| NetProtect | 88 (84, 91) | 67 (61, 72) | 40 (34, 45) | 2.6<br>(2.3, 2.8) | 0.70<br>(0.62, 0.77) | 1.2<br>(1.1, 1.4) |
| p-value |  |  |  |  | p = 0.001 |  |

† Details of the analysis are located in Table S3 Number at risk (functional survival)

### Economic Analysis

Simulation results show that the expected Equivalent Annual Cost (EAC) in $ USD of the three LLINs in the study varied between $1·2 (1.1-1.4) for PermaNet and NetProtect and $1·5 (1.3-1.7) for Olyset, assuming that each net was priced identically at $3·0 (Table 1). The longer-lived nets were approximately 20% lower in EAC as compared to the shorter-lived Olyset product.

### Components of functional survival

#### Attrition

There were significant differences in attrition between net products. Olyset nets were lost at a faster rate than PermaNet 2.0 and NetProtect (Table 2, Table S3). After three years, 55% of Olyset nets were no longer present in households compared to 42% of PermaNet 2.0 and 46% of NetProtect (p<0·001; Table 2). Of the 10,571 nets distributed, 4,964 (46%) were lost to follow up over the whole study period (Table S4).

**Table 2:** Percentage attrition (defined as net loss due to discarding or alternative use of nets) with 95% confidence intervals in parenthesis and hazard ratios after 36 months by net product and time point^†^

| Net product | % Attrition (95% CI) |  |  | Hazard ratio |
| --- | --- | --- | --- | --- |
|  | 10 months<br>n=8,269 | 22 months<br>n=6,324 | 36 months<br>n=3,942 |  |
| Olyset | 7 (5, 8) | 25 (21, 29) | 55 (49, 61) | 1 |
| PermaNet | 5 (3, 6) | 20 (17, 24) | 42 (38, 46) | 0.71 (0.64, 0.79) |
| NetProtect | 6 (4, 8) | 22 (18, 26) | 46 (43, 50) | 0.81 (0.71, 0.93) |
|  |  |  |  | p < 0.001 |

† Details of the analysis are located in Table S3 Number at risk (attrition)

#### Physical integrity

The condition of nets that remained in households deteriorated over the course of the study. At each time point, Olyset had the largest proportion and NetProtect had the smallest proportion of ‘too torn’ nets (Figure 1). The median hole surface area in Olyset increased from 38 cm^2^ at 10 months to 459 cm^2^ after 36 months, compared to 6 cm^2^ to 295 cm^2^ for PermaNet 2.0 and 8 cm^2^ and 152 cm^2^ for NetProtect (Table S5). Questionnaire data showed that in year 3, 70% of nets no longer in use had been discarded when they were perceived as too damaged to be useful. Others were given away (17%), stolen (3%) or repurposed (3%).

**Figure 1:**
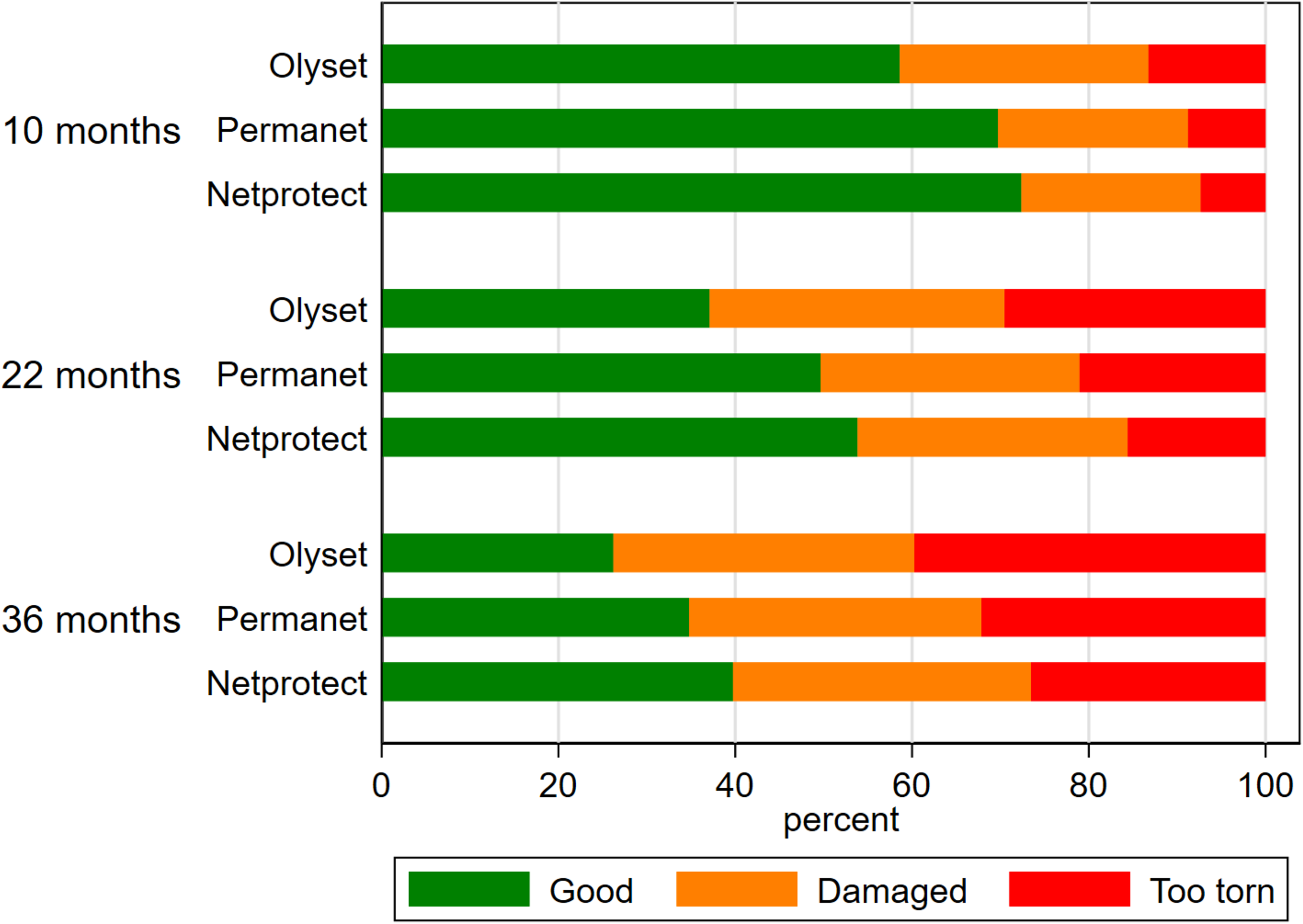
Physical condition of LLINs according to WHO categorisation using proportional Hole Index4 by the three net products and time points. Green shows nets in percentage of nets in good condition (pHI < 65), orange shows % nets in a damaged condition (pHI: 65-364) and red shows % of nets defined as “too torn” (pHI > 364).

#### Bioefficacy

At baseline, all products met optimal WHO bioefficacy criteria. After field use, there were significant differences between the bioefficacy of the net products measured using standard WHO cone and tunnel tests over time (Table 3). At 10 months, 100% of NetProtect and PermaNet 2.0 nets met WHO optimal bioefficacy criteria, compared to 73% of Olyset nets (p<0·001). Nets decreased in bioefficacy through time but even after three years, 96% of NetProtect, 85% of PermaNet 2.0 and 75% of Olyset met WHO criteria for bioefficacy (p=0·017; Table 3).

**Table 3:** Percentages of net products meeting optimal WHO bioefficacy criteria by time point in months. 95% confidence intervals in parenthesis. Numbers passing / numbers tested in square brackets [n/N]^†^.

|  | WHO cone test |  |  | WHO tunnel test |  |  | Overall (cone + tunnel) |  |  |
| --- | --- | --- | --- | --- | --- | --- | --- | --- | --- |
| months | 10 | 22 | 36 | 10 | 22 | 36 | 10 | 22 | 36 |
| Olyset | 4 (1, 14)<br>[2/48] | 8 (2, 20)<br>[4/48] | 14 (5, 27)<br>[6/44] | 72 (57, 84)<br>[33/46] | 78 (62, 89)<br>[34/44] | 71 (54, 85)<br>[27/38] | 73 (58, 85)<br>[35/48] | 79 (65, 90)<br>[38/48] | 75 (60, 87)<br>[33/44] |
| PermaNet | 98 (89, 100)<br>[46/47] | 92 (80, 98)<br>[44/48] | 73 (58, 85)<br>[35/48] | 100 (3, 100)<br>[1/1] | 50 (7, 93)<br>[2/4] | 46 (19, 75)<br>[6/13] | 100 (92, 100)<br>[47/47] | 96 (85, 99)<br>[46/48] | 85 (72, 94)<br>[41/48] |
| NetProtect | 100 (92, 100)<br>[47/47] | 100 (93, 100)<br>[48/48] | 73 (58, 85)<br>[35/48] | n/a | n/a | 85 (55, 98)<br>[11/13] | 100 (92, 100)<br>[47/47] | 100 (93, 100)<br>[48/48] | 96 (86, 99)<br>[46/48] |
| p-value |  |  |  |  |  |  | <0.001 | <0.001 | 0.017 |

<sup>†</sup>Nets are tested by cone test and those that fail WHO optimal insecticide effectiveness criteria of 95% knockdown after 60 minutes or 80% 24-hour mortality are then further tested by tunnel test. Optimal criteria for tunnel test are 80% 24-hour mortality or 90% blood feeding

When whole nets were tested using IACT, 88% of Olyset, 96% of PermaNet 2.0 and 92% of NetProtect passed WHO optimal criteria of 80% mortality and 90% blood feeding inhibition after 3 years. There were differences between products in 24-hour mortality. Olyset showed lower mortality (p<0·001), but all three products showed similar levels of feeding inhibition (Figure 2, Table S6). Mosquito mortality was higher for nets defined as “too torn” (OR = 0·65 (0·49, 0·88), p=0·005), but the differences between the net products remained significant after adjusting for physical condition. Similarly, protection from mosquito bites (feeding inhibition) was considerably lower in nets that were “too torn” (OR = 0·12 (0·08, 0·18), p<0·001), but the differences between the net products remained non-significant after adjusting for physical condition.

**Figure 2:**
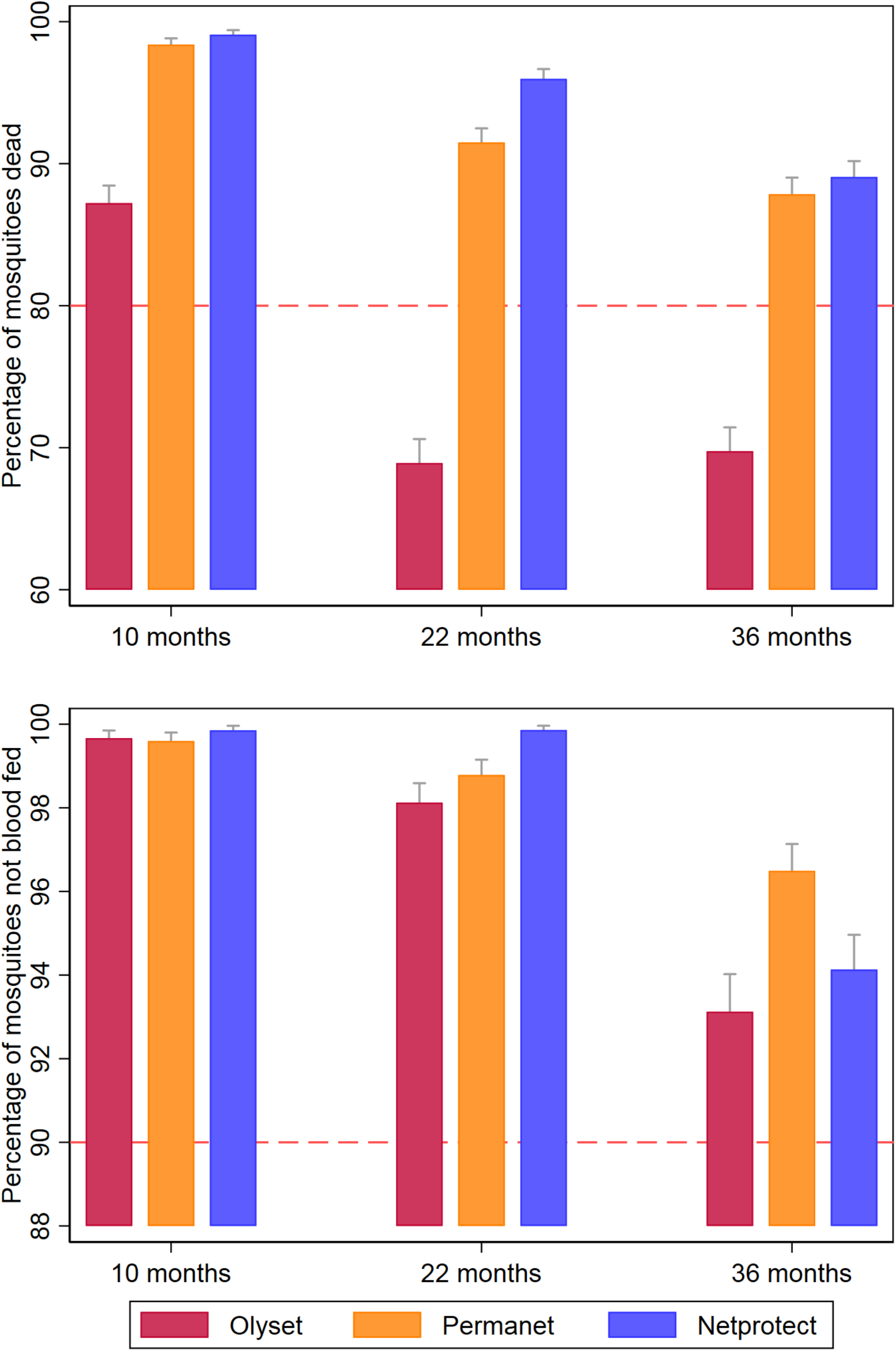
Ifakara-Ambient Chamber Test (IACT) results on mosquito mortality (top panel) and blood feeding inhibition (bottom panel) by net product (red = Olyset; orange = PermaNet; blue = NetProtect) and time point. Optimal WHO criteria (80% mortality; 90% blood feeding inhibition) are indicated by the dashed line.

#### Active ingredient content

At baseline, 100% (10) of Olyset and PermaNet 2.0 and 50% (5) of NetProtect samples complied with their target doses of active ingredient (Table S7). Deltamethrin content lower than the target dose in NetProtect was explained by a high *R*-alpha isomer content (0·35 g/kg on average, 26% of the deltamethrin content), a non-relevant impurity of deltamethrin, which may be formed during the manufacturing process.

After each timepoint, mean permethrin content in Olyset decreased to 16·2 g/kg, 14·8 g/kg and 13·0 g/kg, corresponding to a loss of 20%, 27% and 36% of the original dose, respectively. Mean deltamethrin content of PermaNet 2.0 decreased to 0·75 g/kg, 0·47 g/kg and 0·40 g/kg, corresponding to a loss of 48%, 68% and 72% of the original dose, respectively. Mean deltamethrin content of NetProtect decreased to 0·91 g/kg, 0·52 g/kg and 0·40 g/kg, corresponding to a loss of 33%, 61% and 70% of the original dose, respectively (Table S7).

## Discussion

This large and well-powered study clearly disproves the assumption that all pyrethroid treated LLIN products have similar lifespans. Our data also confirms that the median functional life of the LLINs in our study was closer to two years than three years in Tanzania and for the products tested, as previously reported by a systematic review of LLIN retention data in 39 sub-Saharan African countries.^6^

The WHO’s *Guidelines for procuring public health pesticides*^14^ recommends that the procurement should consider “operational cost” rather than unit price, and an appropriate measure to compare value for money of LLINs would be “cost per median year of net life under local conditions”. We measured the relative durability of nets using functional survival estimates, in terms of the equivalent annual cost (EAC). The cost analysis showed approximately 20% lower EAC when a longer-lasting LLIN (PermaNet 2.0 or NetProtect) was chosen over Olyset, assuming prices for the products were identical. The relative increase in price that is acceptable is also much smaller when the lifetime of the standard reference product increases. Thus, the extension of the life of a product is much more valuable if the comparator product is relatively short-lived, as was seen in this study.

LLIN functional life also has important implications for the selection of new products for resistance management that have a higher unit cost. New pyrethroid plus piperonyl butoxide (PBO) nets may not be as durable as standard pyrethroid nets because PBO is lost rapidly from nets during washing, which reduces their efficacy.^15^ However, in Tanzania, PBO nets continued to have superior public health benefit, two years after distribution.^7^ If the median functional survival of pyrethroid LLINs is two years, then PBO nets may remain cost-competitive.

WHO requires LLIN manufacturers to provide data from three longitudinal field evaluations in different ecologies e.g. West Africa, East Africa and Asia that is used to evaluate LLINs for Prequalification (PQ) listing. While it is recognized that durability is context specific, using the WHO methodology outlined^16,17^, it is possible to routinely generate median functional survival estimates and EAC for at least three locations during PQ evaluation, albeit with a more limited sample size than the present study. The EAC may be a useful metric to compare between products, rather than assessing products based simply on a minimum threshold as is current practice. The role of durability data in LLIN procurement has been side-lined and consideration of its importance in vector control by WHO may re-awaken the LLIN market to reward more durable products that will in turn create incentives for investments in technological advances, research and development by ITN manufacturers.^5,17^

Attrition and physical integrity, the two factors that define functional survival of LLINs,^2^ differed significantly between the three net products. Olyset demonstrated more rapid accumulation of damage and more rapid attrition. In the current study and in previous work we demonstrated that most LLINs were discarded because they were perceived as too damaged to offer protection against mosquito bites or malaria.^18^ Further consideration should be given to developing simple tools to allow countries to assess attrition and fabric integrity during routine surveys to inform planning since it is clear that these two outcomes are both most important in predicting LLIN functional life, are highly variable between contexts and are simpler to collect than bioefficacy or chemical content data.

Of those nets still present after three years, 25-40% were categorized as no longer physically serviceable, depending on the brand. However, even after three years, nets remained highly efficacious when tested by bioassays against insecticide-susceptible malaria vectors. Damage actually increased the mortality of mosquitoes that had entered nets through holes and become trapped, as also observed in other studies.^19^ Indeed, torn LLINs continue to provide both individual and community protection from malaria.^20,21^ Our IACT experiments, demonstrated that the three brands were all highly protective, although Olyset killed significantly fewer mosquitoes than PermaNet 2.0 and NetProtect. It is of note that most damage to the nets is on the bottom section from where they are tucked under a mat or mattress. The act of tucking makes these holes inaccessible to mosquitoes even though the net appears as badly damaged to the user.

However, it is a limitation of the presented study that only susceptible mosquitoes were used for bioefficacy testing as pyrethroid resistance is widespread and increases feeding success and reduces mortality of mosquitoes.^19^ Another limitation is the fact that the study was only conducted in Tanzania and LLIN durability does vary by location. Furthermore, this study was conducted on only three brands of LLINs, all of which are treated with pyrethroids. As new LLINs products come on the market especially those with new insecticides e.g. PBO nets it will be imperative to monitor their comparative durability to ensure that the most cost-effective products are procured for malaria control.

It was demonstrated that nets that are still in households, despite holes, are still protective against mosquito bites and continue to kill mosquitoes, providing personal and community protection. However, if nets are discarded, or no longer used because they are perceived as too damaged, then they have no public health benefit at all. While it is possible to encourage users to retain their damaged but still insecticidal nets through behavioural change communication (BCC) a more effective strategy will be to distribute more physically durable LLINs.^22^

LLINs are the largest single item on the global malaria control budget. Most are distributed through mass campaigns.^3^ More durable LLINs would reduce the required frequency of campaigns and thus the operational costs of distribution per person-year of coverage. It is technically feasible to manufacture more durable LLINs. However, this will happen only if buyers consider cost-effectiveness for coverage ^14^ and demonstrate a preference for longer-lasting and better value-for-money products, rather than considering only the unit price.

## Methods

The study has been described in detail previously.^23^ It took place in 8 districts in Tanzania, selected to be representative of national environmental, ecological and epidemiological settings (Figure 3). Within each district, 10 villages were randomly selected, and within each village 45 households were recruited to participate in the study. All households were randomised to receive one of three LLIN brands on a 1:1:1 ratio, stratified by village. The three brands were Olyset^®^ (permethrin incorporated in 150 denier polyethylene; Sumitomo Chemicals, Japan), PermaNet^®^2.0 (deltamethrin coated on 100 denier polyester; Vestergaard Frandsen, Switzerland), or NetProtect^®^ (deltamethrin incorporated into 110 denier polyethylene; BestNet, Denmark). Distribution of study nets took place between October and December 2013. All nets owned by the participating households were collected and replaced with enough nets to cover all sleeping spaces. Before distribution, a sample of ten nets per product was quality tested. Nets were the same size and colour, labelled by a five-digit serial number so that participants and investigators remained blinded to the LLIN product until data collection was complete. In total, 3,393 households were randomised (1,132 to Olyset, 1,127 to PermaNet 2.0 and 1,134 to NetProtect) to which 10,571 nets were distributed (Trial Characteristics Table S1).

**Figure 3:**
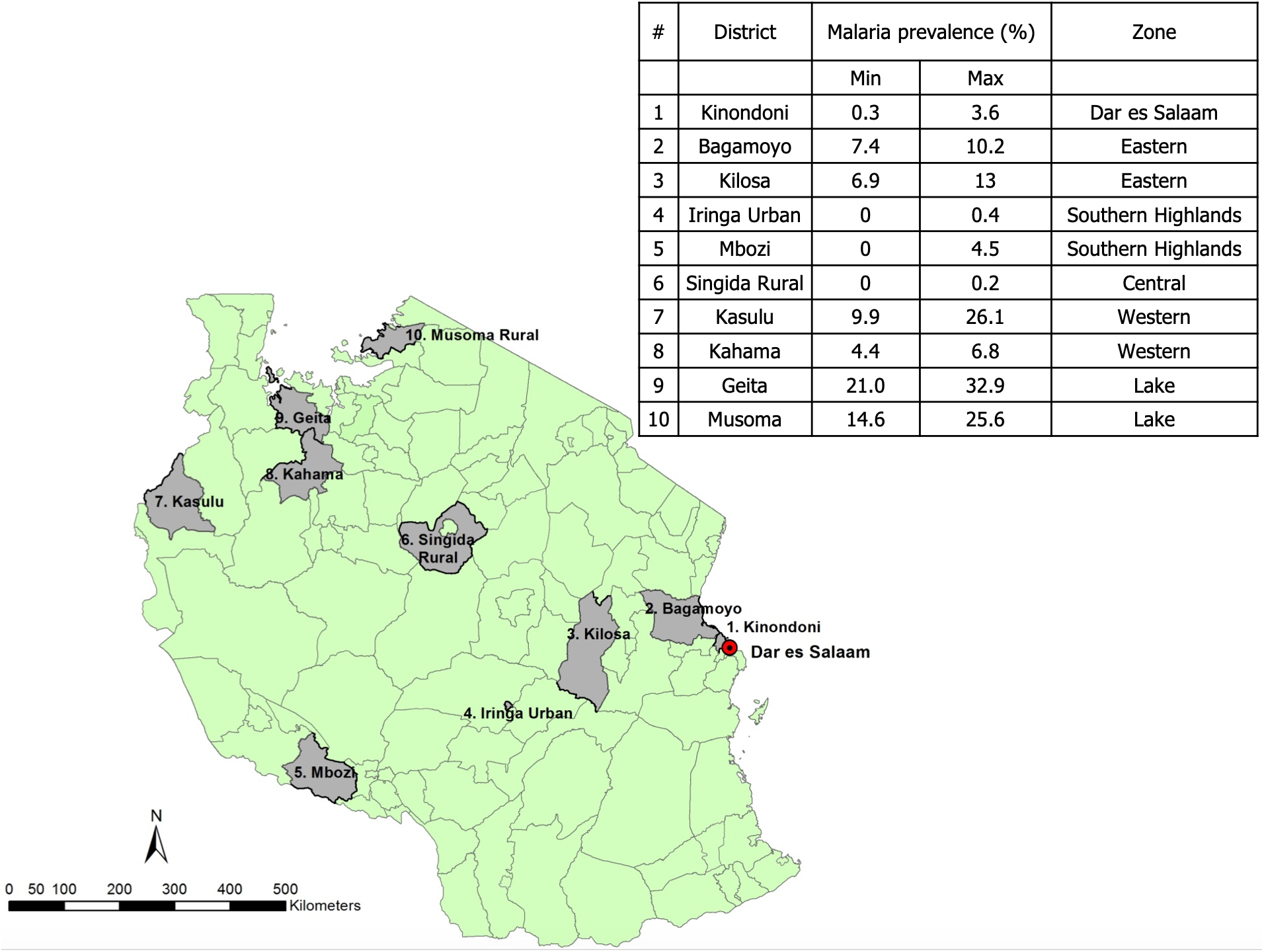
Map of ABCDR study districts with 2015 malaria prevalence data (% of children aged 6-59 months diagnosed with malaria by Rapid Diagnostic Test and microscopy)

Surveys were conducted among all consenting study households when the LLINs were distributed and at three follow up points: 10 months (August – October 2014), 22 months (August – October 2015) and 36 months (October – December 2016) (Study Flow Table S8). Serial numbers of nets, linked to household identifying codes in a master list, enabled follow-up of each net at each time point. At each follow up visit information on each LLIN was collected, including whether the net was present in the house, reasons why it was not present, and whether the net was in use. Physical integrity of LLINs was measured on a random sample of three nets per household by counting the number, location and size of holes.^16,17^ Socio-economic variables and a household member roster were also recorded. Electronic data capture was used for all surveys.

In addition to the data collected as part of the household surveys, at each time point 48 LLINs from each brand were randomly sampled from the master list and returned to the laboratory in Bagamoyo, Tanzania for bioefficacy and chemical analysis using standard WHO methods^16,17^ and additional Ifakara Ambient Chamber Test (IACT).^24^ Table 4 describes the different components of LLIN durability, the test conducted to obtain the data, the outcome indicators for statistical analysis and the corresponding WHO threshold criteria.^4,16,17^ The numbers of LLINs tested for each of the components of LLIN durability are listed in Table S8.

**Table 4.** LLIN durability components.

| Component | Definition | Test conducted | Outcome indicators | WHO criteria or industry standard |
| --- | --- | --- | --- | --- |
| **Attrition** | Net loss from household through discarding or use for alternative purpose‡ | Household survey | Net presence |  |
| **Physical integrity** | Physical state of the net to estimate bite protection | Count number, location and size of hole(s) of max. 3 nets per household | Holed surface area measured by the proportionate Hole Index (pHI)<sup>4†</sup><br>Median hole surface area (cm<sup>2</sup>) | pHI 0-64: good<br>pHI 65-642: damaged<br>pHI<642: serviceable<br>pHI>643: too torn/unserviceable |
| **Functional survival<sup>4</sup>** | Estimation of nets still in households in serviceable condition | Median survival analysis | (Number of nets present and serviceable) / (Number of nets originally received and not given away or lost to follow up) | Median net survival in years = time point at which the estimate of functional survival crosses 50% |
| **Biological efficacy** | Ability of net to incapacitate or kill susceptible anopheline mosquitoes after contact with insecticide | Ifakara Ambient Chamber Test (IACT)*: whole nets<sup>17</sup> | Proportion of mosquitoes dead at 24 hours<br>Proportion of mosquitoes not blood fed |  |
|  |  | WHO cone/tunnel test: 25x25cm pieces<sup>15</sup> | Net samples meeting optimal bioefficacy criteria | 1hr Knock-Down ≥ 95%<br>or<br>24hr Mortality ≥ 80% or<br>Blood feeding Inhibition ≥ 90% |
| **Insecticide content** | Amount of active ingredient in the net | Permethrin: Gas Chromatography with Flame Ionisation Detection (GC-FID)<br>Deltamethrin: High Performance Liquid Chromatography with UV Diode Array Detection (HPLC-DAD) | Compliance of nets with WHO specifications at baseline<br>Loss of active ingredient over time | Olyset: 20 g/kg ± 25% [15 -25 g/kg]<br>PermaNet: 1.4 g/kg ± 25% [1.05-1.75 g/kg]<br>NetProtect: 1.8 g/kg ± 25% [1.35 -2.25 g/kg] |

‡ Nets that were reported as given away, sold or stolen were not in the denominator (lost to follow up).
† proportionate Hole Index (pHI): Number of holes of each size category measured in the field are weighted by the approximate surface area of the holes to provide a single measure of damage per net.
\*previously described as Ifakara Tunnel Test (ITT)<sup>14</sup>

First, the protective efficacy of whole nets returned from the field was evaluated using IACT.^24^ Each night, ten male volunteers slept underneath one of the nets (or an untreated control net to monitor the quality of the bioassay) between 21.00 – 6.00hrs in a small chamber similar in size to a bedroom, within a screened compartment. At 21.00hrs, 30 laboratory-reared mosquitoes were released into the chamber. The next morning, all mosquitoes within the compartment were recaptured, and scored for 24-hour mortality and blood-feeding inhibition. Each LLIN was tested twice on two consecutive nights. Subsequently, net pieces (25×25cm^2^) were cut following the WHO sampling pattern and standard WHO cone bioassays were carried out.^16^ If nets did not meet WHO optimal bioefficacy criteria for cone tests (Table 4), WHO tunnel tests were conducted.^16^ Insecticide content analyses were performed using standard CIPAC methods for LLIN insecticide content (Olyset: 331/LN/M/3, PermaNet 2.0 333/LN/(M)/3, NetProtect 333/LN/(M2)/3). All mosquito assays were conducted with fully pyrethroid-susceptible 2-8-day old nulliparous female *Anopheles gambiae* sensu stricto (Ifakara strain).

### Statistical Analysis

All statistical analyses were conducted using Stata Statistical Software: Release 13 (StataCorp LP, TX). Attrition and functional survival (Table 4) were calculated using Kaplan-Meier estimators. For both endpoints, nets reported as given away, sold or stolen were treated as lost to follow up. Hazard ratios for the difference in attrition and functional survival were calculated using discrete time survival analysis using a complementary log-log model.^25^ Robust standard errors were used to account for the highest level of clustering (district).^26^ Of nets that were present, net condition was defined, following WHO recommendations, as either being “good”, “damaged” (combined to be “serviceable”), or “too torn/unserviceable” (Table 4). Negative binomial regression was used to compare hole surface area between net products. A Chi-squared test assessed the proportion of nets of each product passing the WHO bioefficacy criteria based on combined cone and tunnel tests, adjusted for control mortality. Logistic regression was used to analyse mortality and blood feeding inhibition from the IACT test; results were adjusted for chamber and experimental night and robust standard errors were used to take account of nets being tested multiple times.

### Economic Analysis

The equivalent annual cost (EAC) of an LLIN was calculated according to the standard formula.^27^ To assess the value of longer functional survival we used Equation 1 where *b* is the ratio of the lifespan of the more durable product to the lifespan of reference net *n*. The variable *r* is the discount rate. This relationship shows that for any change in net lifespan from *n* to *bn* the relative increase in price, *a*, which would yield an identical EAC for the two products. Other factors being equal a relative price increase less than *a* would favour the new, longer-lasting LLIN, while relative price increases greater than *a* would favour the standard reference net.

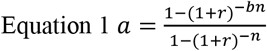

Simulation of EACs for products tested in the study was conducted using Monte Carlo methods, assuming a 3% discount rate, as is standard in health economic analysis. The baseline survival function for LLINs was estimated by regressing the survival proportions of Olyset nets derived from Kaplan-Meier analysis (Table S4) against time. The survival function was converted into a baseline hazard and net failure lifetimes were simulated for a cohort of 500 LLINs assuming a Weibull distribution of times to failure (in terms of functional survival). The results of the cohort were summarised by estimating the median lifetime and this process was repeated 10,000 times for each net type, yielding an estimate of the expected median lifetime and quantiles of its expected distribution. Results were converted into EACs with 95% quantiles. Distributional assumptions for the baseline hazard and the parameters of the Weibull distribution were fitted to the results presented in Table 1. The baseline hazard and proportional hazard were simulated with log normal distributions (Table S9).

### Ethics

Ethical approval was granted from ethical review committees at London School of Hygiene & Tropical Medicine (6333/A443), Ifakara Health Institute (IHI/IRB/AMM/ No: 07-2014) and the Tanzanian National Institute for Medical Research (NIMR/HQ/R.8c/Vol. I/285). Community sensitization meetings were held prior to study inception and written informed consent was obtained from the head of the household, or another adult household member of participating households before each survey. Human volunteers for the IACT experiment were all IHI staff members with appropriate training who gave written informed consent.

## Data Availability

The datasets supporting the conclusions of this article are available from the Norwegian Centre for Research Data (NSD)

http://www.nsd.uib.no/nsd/english/index.html

## Acknowledgements

Funding for the project was provided by Research Council of Norway. Our special thanks are addressed to all technical staff at Ifakara health Institute, at Kingani Insectary for conducting data collection in the laboratory and semi field experiments. We thank the LLIN manufacturers Sumitomo Chemical (Olyset), Vestergaard Frandsen (PermaNet) and BestNet (NetProtect) for their donation of the LLINs free of charge. Special thanks to Dr. Karen Kramer and Renate Mandike for their thoughtful comments that helped shape the study design and support at study inception. The research was made possible by the Research Council of Norway through the ABCDR Project no. 220757. Sarah Moore was funded by awards from IVCC and Notre Dame OPP1081737. Neither the sponsor nor the LLIN manufacturers had any role in study design; in the collection, analysis, and interpretation of data; in the writing of the report; and in the decision to submit the paper for publication. The Director-General of the National Institute for Medical Research (NIMR) in Tanzania gave permission to publish this paper.

## Availability of data and materials

The datasets supporting the conclusions of this article are available from the Norwegian Centre for Research Data (NSD), http://www.nsd.uib.no/nsd/english/index.html

## Supplementary material

**Table S1.**
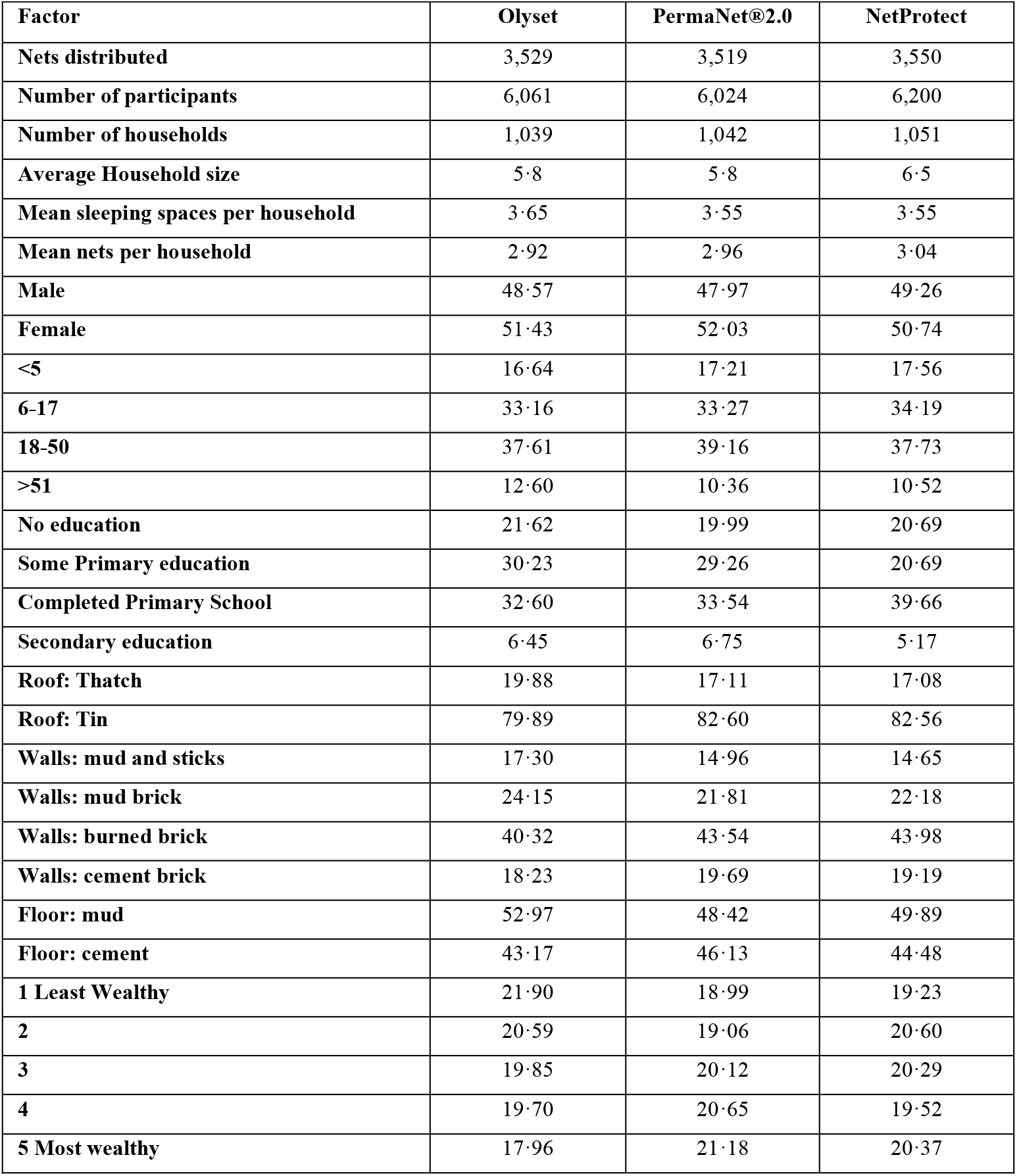
Household and Socioeconomic characteristics of participating households in each study arm.

| Factor | Olyset | PermaNet®2.0 | NetProtect |
| --- | --- | --- | --- |
| Nets distributed | 3,529 | 3,519 | 3,550 |
| Number of participants | 6,061 | 6,024 | 6,200 |
| Number of households | 1,039 | 1,042 | 1,051 |
| Average Household size | 5·8 | 5·8 | 6·5 |
| Mean sleeping spaces per household | 3·65 | 3·55 | 3·55 |
| Mean nets per household | 2·92 | 2·96 | 3·04 |
| Male | 48·57 | 47·97 | 49·26 |
| Female | 51·43 | 52·03 | 50·74 |
| <5 | 16·64 | 17·21 | 17·56 |
| 6-17 | 33·16 | 33·27 | 34·19 |
| 18-50 | 37·61 | 39·16 | 37·73 |
| >51 | 12·60 | 10·36 | 10·52 |
| No education | 21·62 | 19·99 | 20·69 |
| Some Primary education | 30·23 | 29·26 | 20·69 |
| Completed Primary School | 32·60 | 33·54 | 39·66 |
| Secondary education | 6·45 | 6·75 | 5·17 |
| Roof: Thatch | 19·88 | 17·11 | 17·08 |
| Roof: Tin | 79·89 | 82·60 | 82·56 |
| Walls: mud and sticks | 17·30 | 14·96 | 14·65 |
| Walls: mud brick | 24·15 | 21·81 | 22·18 |
| Walls: burned brick | 40·32 | 43·54 | 43·98 |
| Walls: cement brick | 18·23 | 19·69 | 19·19 |
| Floor: mud | 52·97 | 48·42 | 49·89 |
| Floor: cement | 43·17 | 46·13 | 44·48 |
| 1 Least Wealthy | 21·90 | 18·99 | 19·23 |
| 2 | 20·59 | 19·06 | 20·60 |
| 3 | 19·85 | 20·12 | 20·29 |
| 4 | 19·70 | 20·65 | 19·52 |
| 5 Most wealthy | 17·96 | 21·18 | 20·37 |

**Table S2:** Reported net use the previous night by net product and time point. Data represent numbers of respondents (percent) reporting use of nets.

|  | Olyset | PermaNet | NetProtect | Total | p-value |
| --- | --- | --- | --- | --- | --- |
| **10 months** |  |  |  |  | P = 0·195 |
| **not used** | 440 (20·4) | 410 (18·6) | 448 (20·8) | 1298 (19·9) |  |
| **used** | 1714 (79·6) | 1799 (81·4) | 1705 (79·2) | 5218 (80·1) |  |
| **total** | 2154 | 2209 | 2153 | 6516 |  |
| **22 months** |  |  |  |  | P = 0·648 |
| **not used** | 514 (32·2) | 518 (29·8) | 538 (32·4) | 1570 (31·4) |  |
| **used** | 1082 (67·8) | 1221 (70·2) | 1122 (67·6) | 3425 (68·6) |  |
| **total** | 1596 | 1739 | 1660 | 4995 |  |
| **36 months** |  |  |  |  | P = 0·189 |
| **not used** | 358 (49·3) | 449 (45·2) | 471 (52·5) | 1278 (48·8) |  |
| **used** | 368 (50·7) | 545 (54·8) | 426 (47·5) | 1339 (51·2) |  |
| **total** | 726 | 994 | 897 | 2617 |  |

**Table S3.** Number at risk (attrition)

|  | Time point (months) | At risk | censored | Failed |
| --- | --- | --- | --- | --- |
| **Olyset** | 10 | 3520 | 741 | 187 |
|  | 22 | 2592 | 501 | 404 |
|  | 36 | 1687 | 406 | 510 |
| **PermaNet 2.0** | 10 | 3513 | 759 | 132 |
|  | 22 | 2622 | 444 | 351 |
|  | 36 | 1827 | 439 | 383 |
| **NetProtect** | 10 | 3538 | 758 | 163 |
|  | 22 | 2617 | 504 | 367 |
|  | 36 | 1746 | 412 | 413 |

**Table S4.** Number at risk (functional survival)

|  | Time point (months) | At risk | censored | Failed |
| --- | --- | --- | --- | --- |
| **Olyset** | 10 | 3520 | 1019 | 451 |
|  | 22 | 2050 | 399 | 554 |
|  | 36 | 1097 | 307 | 398 |
| **PermaNet** | 10 | 3513 | 1002 | 314 |
|  | 22 | 2197 | 406 | 470 |
|  | 36 | 1321 | 331 | 404 |
| **NetProtect** | 10 | 3538 | 1020 | 307 |
|  | 22 | 2211 | 407 | 436 |
|  | 36 | 1368 | 372 | 403 |

**Table S5:** Median hole surface area in cm^2^ and interquartile range (IQR) by net product and time point.

|  |  | Olyset | PermaNet | NetProtect | Total | p-value |
| --- | --- | --- | --- | --- | --- | --- |
| Median hole surface area in cm<sup>2</sup> (IQR) | 10 months | 38 (0 - 308) | 6 (0 - 145) | 8 (0 - 94) | 19 (0 - 182) | P < 0.001 |
|  | 22 months | 247 (13 - 969) | 84 (0 - 614) | 61 (0 - 364) | 97 (1 - 669) | P < 0.001 |
|  | 36 months | 459 (66 - 1708) | 295 (29 - 1220) | 152 (13 - 838) | 277 (31 - 1185) | P < 0.001 |

**Table S6:**
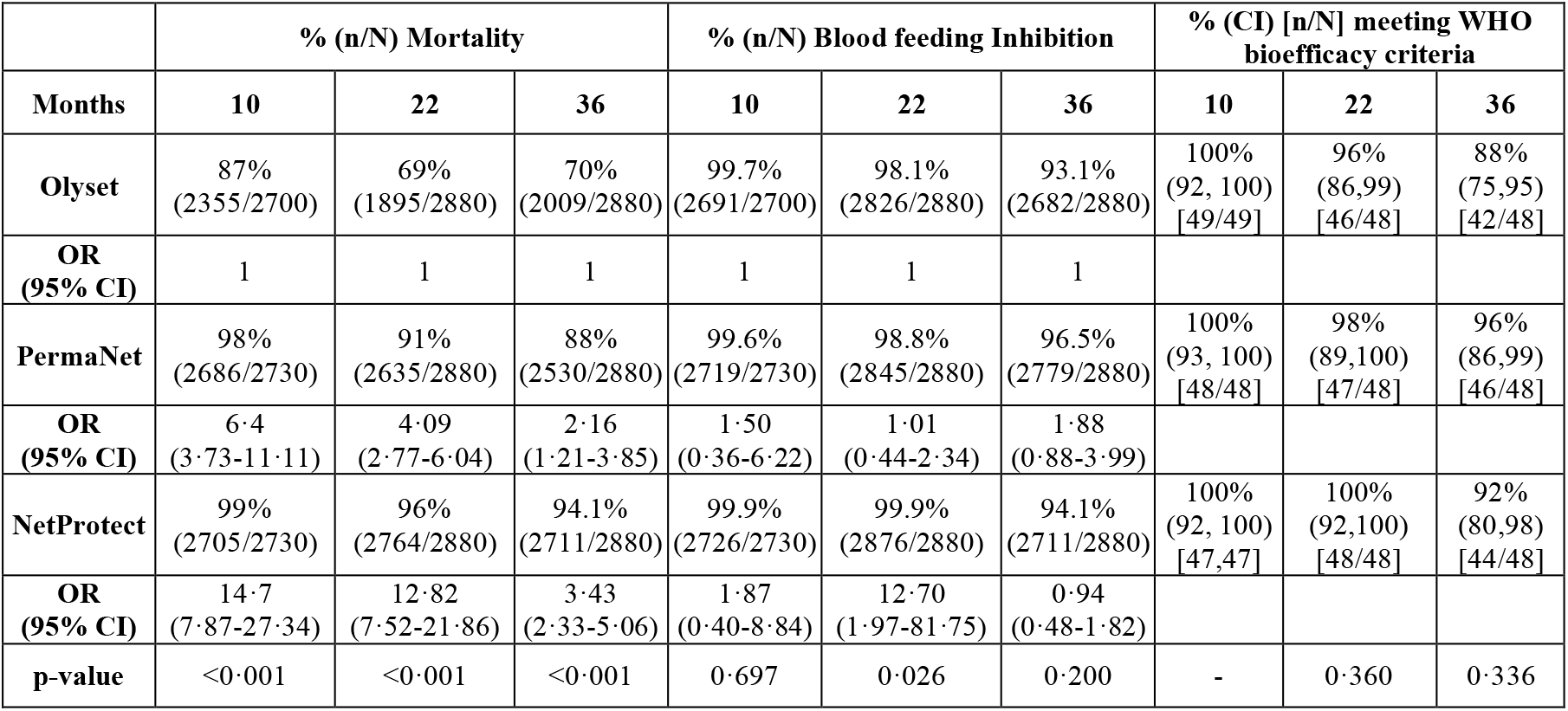
IACT test results on mosquito mortality and blood feeding inhibition by net product and time point in months. Data were analysed by binary logistic regression and the Odds Ratio (OR) and 95% confidence interval (95% CI) of the Odds Ratio are shown.

|  | % (n/N) Mortality |  |  | % (n/N) Blood feeding Inhibition |  |  | % (CI) [n/N] meeting WHO bioefficacy criteria |  |  |
| --- | --- | --- | --- | --- | --- | --- | --- | --- | --- |
| Months | 10 | 22 | 36 | 10 | 22 | 36 | 10 | 22 | 36 |
| Olyset | 87%<br>(2355/2700) | 69%<br>(1895/2880) | 70%<br>(2009/2880) | 99.7%<br>(2691/2700) | 98.1%<br>(2826/2880) | 93.1%<br>(2682/2880) | 100%<br>(92, 100)<br>[49/49] | 96%<br>(86,99)<br>[46/48] | 88%<br>(75,95)<br>[42/48] |
| OR (95% CI) | 1 | 1 | 1 | 1 | 1 | 1 |  |  |  |
| PermaNet | 98%<br>(2686/2730) | 91%<br>(2635/2880) | 88%<br>(2530/2880) | 99.6%<br>(2719/2730) | 98.8%<br>(2845/2880) | 96.5%<br>(2779/2880) | 100%<br>(93, 100)<br>[48/48] | 98%<br>(89,100)<br>[47/48] | 96%<br>(86,99)<br>[46/48] |
| OR (95% CI) | 6.4<br>(3.73-11.11) | 4.09<br>(2.77-6.04) | 2.16<br>(1.21-3.85) | 1.50<br>(0.36-6.22) | 1.01<br>(0.44-2.34) | 1.88<br>(0.88-3.99) |  |  |  |
| NetProtect | 99%<br>(2705/2730) | 96%<br>(2764/2880) | 94.1%<br>(2711/2880) | 99.9%<br>(2726/2730) | 99.9%<br>(2876/2880) | 94.1%<br>(2711/2880) | 100%<br>(92, 100)<br>[47,47] | 100%<br>(92,100)<br>[48/48] | 92%<br>(80,98)<br>[44/48] |
| OR (95% CI) | 14.7<br>(7.87-27.34) | 12.82<br>(7.52-21.86) | 3.43<br>(2.33-5.06) | 1.87<br>(0.40-8.84) | 12.70<br>(1.97-81.75) | 0.94<br>(0.48-1.82) |  |  |  |
| p-value | <0.001 | <0.001 | <0.001 | 0.697 | 0.026 | 0.200 | - | 0.360 | 0.336 |

**Table S7:** Number of nets, mean active ingredient (AI) content (g/kg), range (g/kg) and between net variation (%RSD), percentage of active ingredient lost over time, mean *R*-alpha isomer content (g/kg) and percentage of deltamethrin (only for PermaNet 2.0 and NetProtect), in net samples at baseline and three follow up time points.

| LLIN | Test | Months after distribution |  |  |  |
| --- | --- | --- | --- | --- | --- |
|  |  | 0 | 10 | 22 | 36 |
| Olyset | Number | 10 | 49 | 48 | 48 |
|  | AI content (mean) | 20.3 | 16.2 | 14.8 | 13.0 |
|  | AI content (range) | 20.0 – 20.9 | 8.4 – 19.3 | 7.5 – 20.0 | 3.3 – 19.8 |
|  | AI variation (RSD) | 1% | 13% | 19% | 33% |
|  | AI lost | - | 20% | 27% | 36% |
| PermaNet 2.0 | Number | 10 | 48 | 48 | 48 |
|  | AI content (mean) | 1.45 | 0.75 | 0.47 | 0.40 |
|  | AI content (range) | 1.36 – 1.68 | 0.03 – 1.83 | < 0.01 – 1.51 | < 0.01 – 1.55 |
|  | AI variation (RSD) | 7% | 58% | 77% | 106% |
|  | AI lost | - | 48% | 68% | 72% |
|  | R-alpha content (mean) | 0.02 | < 0.01 | < 0.01 | < 0.01 |
|  | R-alpha (% of deltamethrin) | 1.4% | < 1.3% | < 1.3% | < 1.3% |
| NetProtect | Number | 10 | 47 | 48 | 48 |
|  | AI content (mean) | 1.35 | 0.91 | 0.52 | 0.40 |
|  | AI content (range) | 1.26 – 1.43 | 0.07 – 1.88 | 0.19 – 1.24 | 0.05 – 0.99 |
|  | AI variation (RSD) | 4% | 32% | 42% | 60% |
|  | AI lost | - | 33% | 61% | 70% |
|  | R-alpha content (mean) | 0.35 | 0.29 | 0.25 | 0.20 |
|  | R-alpha (% of deltamethrin) | 26% | 32% | 48% | 50% |

**Table S8.** Study Flow.

|  | 2014 (Year 1) | 2015 (Year 2) | 2016 (Year 3) |
| --- | --- | --- | --- |
| Households interviewed | 87.2% (2,962/3,398) | 95.7% (2,834/2,962) | 96.3% (2,730/2,834) |
| HH loss to follow up | 12.8% (436/3,398) | 4.3% (128/2,962) | 3.7% (104/2,834) |
| ABCDR nets loss to follow up | 8.3% (880/10,598) | 7.2% (539/7,477) | 10.4% (551/5,311) |
| Nets lost from households | 23.1% (2,241/9,726) | 23.5% (1,627/6,938) | 39.0% (1,855/4,760) |
| Nets inspected for holes | 82.5% (6,166/7,477) | 90.1% (4,783/5,311) | 86.7% (2,519/2,905) |
| Nets evaluated by IACT | 142 | 144 | 140 |
| Nets evaluated for bioefficacy | 142 | 144 | 140 |
| Nets evaluated for chemical content | 144 | 144 | 144 |

**Table S9.**
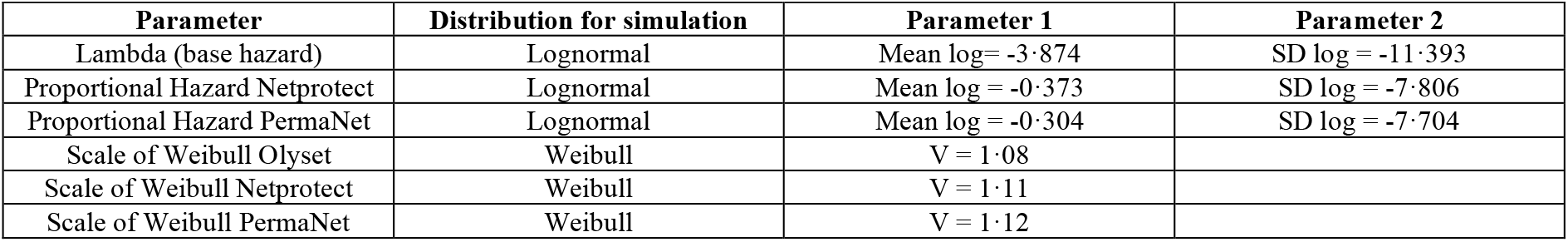
Parameters used in simulation of lifetimes for equivalent annual cost simulation analysis.

